# Insights into the molecular mechanism of anticancer drug ruxolitinib repurposable in COVID-19 therapy

**DOI:** 10.1101/2020.12.29.20248986

**Authors:** Manisha Mandal, Shyamapada Mandal

## Abstract

Due to non-availability of specific therapeutics against COVID-19, repurposing of approved drugs is a reasonable option. Cytokines imbalance in COVID-19 resembles cancer; exploration of anti-inflammatory agents, might reduce COVID-19 mortality. The current study investigates the effect of ruxolitinib treatment in SARS-CoV-2 infected alveolar cells compared to the uninfected one from the GSE5147507 dataset. The protein-protein interaction network, biological process and functional enrichment of differentially expressed genes were studied using STRING App of the Cytoscape software and R programming tools. The present study indicated that ruxolitinib treatment elicited similar response equivalent to that of SARS-CoV-2 uninfected situation by inducing defense response in host against virus infection by RLR and NOD like receptor pathways. Further, the effect of ruxolitinib in SARS-CoV-2 infection was mainly caused by significant suppression of IFIH1, IRF7 and MX1 genes as well as inhibition of DDX58/IFIH1-mediated induction of interferon-I and -II signalling.

## Introduction

The SARS-CoV-2 infection induced hyperinflammatory state is a general manifestation in severe and critical COVID-19 leading to significant morbidity and mortality worldwide (Yeleswaram et al. 2020). There is an intense need of repurposing the approved available drugs for COVID-19 treatment due to non-availability of specific therapeutics (Mandal and Mandal, 2000). Ruxolitinib, (3R)-3-cyclopentyl-3-[4-(7H-pyrrolo[2,3-d]pyrimidin-4-yl)-1H-pyrazol-1-yl] propanenitrile, an FDA approved antineoplastic agent, is one such drug repurposable in COVID-19, that acts as Janus Kinase (JAK) 1 and JAK2 inhibitors to suppress the release of proinflammatory cytokines IL1, IL6, IL8, IL12, TNFA, INFG, VEGF, TGFB, FGF, PDGF, GMCSF, GCSF (Lussana and Rambaldi 2017). It is used in the treatment of polycythemia vera, myelofibrosis of the primary, post-polycythemia vera and post-essential thrombocythemia types as well as steroid-refractory acute graft-versus-host disease post allogeneic stem cell transplantation (https://www.accessdata.fda.gov/drugsatfda_docs/label/2019/202192s017lbl.pdf). Notably, JAK1 and JAK2 are important regulators of hematopoietic homeostasis and immune system development through downstream target gene (STAT3/5, AKT, and ERK) phosphorylation, thereby stimulating cytokines and chemokines production (Ajayi et al. 2018).

There have been reports on the effect of ruxolitinib treatment in COVID-19 patients ameliorating the symptoms, improving the radiographic findings, blood biochemical parameters and immune cell status. Administration of ruxolitinib in severe COVID-19 patients demonstrated improved chest CT, faster recovery from lymphopenia, alleviation of exuberant proinflammatory cytokine (IL1, IL2, IL4, IL6, IL10, IL12, IL13, IL17, GMCSF, GCSF, IP10, MCP1, MIP1A, HGF, INFG, TNFA) (Cao et al. (2020), Neubauer et al. (2020), Rosee et al. (2020), https://clinicaltrials.gov/ct2/show/NCT04348071) along with mitigation of plasma ferritin concentration, soluble IL2R levels, and STAT-phosphorylation, STAT1 dependent CD8+ T cell proliferation (Zhong et al. 2020 Lancet Rheumatol), reduction of cytotoxic T-lymphocytes and increment of T_reg_ cells (https://clinicaltrials.gov/ct2/show/NCT04334044). However, the key molecular interactions based on differentially expressed genes (DEGs) in SARS-COV-2 infection with or without ruxolitinib treatment, in repurposing the drug for COVID-19 treatment, have not been widely explored.

Therefore, the current study is an attempt to investigate the effect of ruxolitinib treatment in SARS-CoV-2 infected alveolar cells compared to the uninfected one from the GSE5147507 dataset, emphasizing the protein-protein interaction (PPI) network, biological process and functional pathways enrichment associated with the DEGs. To achieve this, STRING App of the Cytoscape software, version 3.7.2 (https://cytoscape.org/) was applied along with R programming (https://www.r-project.org/; version 3.6.1 [2019-07-05]) gplots and ggplot tools from the Biobase packages (https://CRAN.R-project.org/package=gplots) of the Bioconductor project (Huber et al., 2015).

## Methods

The gene expression dataset GSE5147507 was obtained from the Gene Expression Omnibus (GEO) database (www.ncbi.nlm.nih.gov/geo/) related to transcriptional response to SARS-CoV-2 infection (Blanco-Melo et al., 2020), from where samples were selected containing independent biological triplicates of (i) transformed lung alveolar (A549) cells transduced with a vector expressing human ACE2 mock treated (GSM4486157: Series16_A549-ACE2_Mock_1, GSM4486158:Series16_A549-ACE2_Mock_2, and GSM4486159:Series16_A549-ACE2_Mock_3), (ii) ACE2 transduced A549 cells with SARS-CoV-2 infection and without ruxolitinib treatment (GSM448616:Series16_A549-ACE2_SARS-CoV-2_1, GSM4486161:Series16_A549-ACE2_SARS-CoV-2_2, and GSM4486162:Series16_A549-ACE2_SARS-CoV-2_3), and (iii) ACE2 transduced A549 cells with SARS-CoV-2 infection and ruxolitinib (500 nM) treatment (GSM4486163:Series16_A549-ACE2_SARS-CoV-2_Rux_1, GSM4486164:Series16_A549-ACE2_SARS-CoV-2_Rux_2, GSM4486165:Series16_A549-ACE2_SARS-CoV-2_Rux_3).

The differentially expressed genes (DEGs) were identified in (i) transformed lung alveolar (A549) cells transduced with ACE2 and infected with SARS-CoV-2 compared to the mock treated cell line, (ii) ACE2-induced A549 cell line infected with SARS-CoV-2 and treated with ruxolitinib compared to the ruxolitinib untreated cell line with SARS-CoV-2 infection. The threshold for DEGs were set as |log_10_FoldChange| (|log_10_FC|)>1 and p value<0.05. The PPI network was constructed with the DEGs using STRING App of the Cytoscape software, version 3.7.2 (https://cytoscape.org/), as per Shannon et al. (2003). The DEGs |log_10_FC| were analysed at the functional level for GeneOntology (GO) and functional pathway enrichment using the STRING Enrichment App of Cytoscape, with p value<0.05 as the cut-off criterion.

## Results and Discussion

The microarray gene expression following SARS-CoV-2 infection, from GSE5147507 dataset (www.ncbi.nlm.nih.gov/geo/) submitted by Blanco-Melo et al. (2020), was analyzed using bioinformatics approaches, to identify the transcriptional signatures in COVID-19 pathogenesis by gene interaction networks, gene ontology and functional pathway enrichment analysis in SARS-CoV-2 infection with or without ruxolitinib pre-treatment compared to that without infection. A total of 21797 genes were obtained in the microarray expression from ACE2 induced transformed alveolar cell lines comprising triplicate samples, each of mock treated (without SARS-CoV-2 infection and ruxolitinib treatment), SARS-CoV-2 infected, and SARS-CoV-2 infected treated with ruxolitinib.

Analysis of the DEGs between the mock treated and SARS-CoV-2 infected cell line yielded 107 upregulated and 139 downregulated genes (log_10_FC>1), while the SARS-CoV-2 infected cell line having ruxolitinib treatment expressed significantly enriched downregulated genes (log_10_FC>1) (n=44) only compared to the infected but untreated cell line. The SARS-CoV-2 infected cells treated with ruxolitinib demonstrated similar gene expression as found in the uninfected cell line, while the SARS-CoV-2 infected cells overexpressed the genes related to the defense response in host to virus infection in the order: IFIT1>IFIT3>IFI6>SAMD9>MX1>OASL>HERC5>IFIH1>ISG15>OAS2>NLRC5, in terms of microarray signal intensities averaged over samples (range 942-9716) (Figure 1a), implying that ruxolitinib showed plausible potentiality of ameliorating COVID-19 condition. Ruxolitinib treatment elicited downregulated DEG response amongst which the key candidate genes were UBA7 with highest log_10_FC value −1.72, and IFIH1, IRF7 and MX1 with maximum degree connectivity of 27 (Figure 1b). The UBA7 (Ubiquitin Like Modifier Activating Enzyme 7) is a ubiquitin-protein transferase and ubiquitin activating enzyme involved in MHC-I mediated antigen processing, presentation and innate immune system related to acute promyelocyte leukemia and lung cancer (GeneCards, 2020). The downregulated DEG possessing highest log_10_FC (−1.34242) was SLC1A2 (Solute Carrier Family 1 Member 2), and the highest degree connectivity bearing DEGs: IRF7 (n=25), IFIH1 (n=23) and MX1 (n=22) were upregulated in SARS-CoV-2 infected cell line compared to uninfected one (Figure 1c). The IFIH1, IRF7 and MX1 were vital players in distinguishing the effect of ruxolitinib treatment in SARS-CoV-2 infection compared to ruxolitinib untreated but SARS-CoV-2 infected cell, which were suppressed in the former and enhanced in the latter (Figure 1b and 1c). The IFIH1 (Interferon Induced with Helicase C Domain 1), a cytoplasmic sensor of viral nucleic acids, associates with mitochondria antiviral signaling protein (MAVS/IPS1) to induce IKK-related kinases: TBK1 and IKBKE, to phosphorylate interferon regulatory factors IRF3 and IRF7 for the activation of antiviral responses including RIG-I/MDA5 mediated type I interferon signaling and proinflammatory cytokines (GeneCards, 2020). The MX1 (MX Dynamin Like GTPase 1) is also induced in type I and type II interferon antiviral responses to inhibit RNA viral replication (GeneCards, 2020). The SARS-CoV-2 infection stimulated antiviral mechanism by DDX58/IFIH1-mediated interferon α/β-signaling involving EGR1, OASL, SOCS1, MX2, OAS2, XAF1, IFIT1, IFIT3, IFIT2, IFNB1, RSAD2, IRF7, MX1, IFITM1, IFNA5 genes, amongst which the major candidate gene upregulated was IFNB1 with highest log_10_FC 2.32, and degree connectivity 21 (Figure 1c).

**Figure 1:**
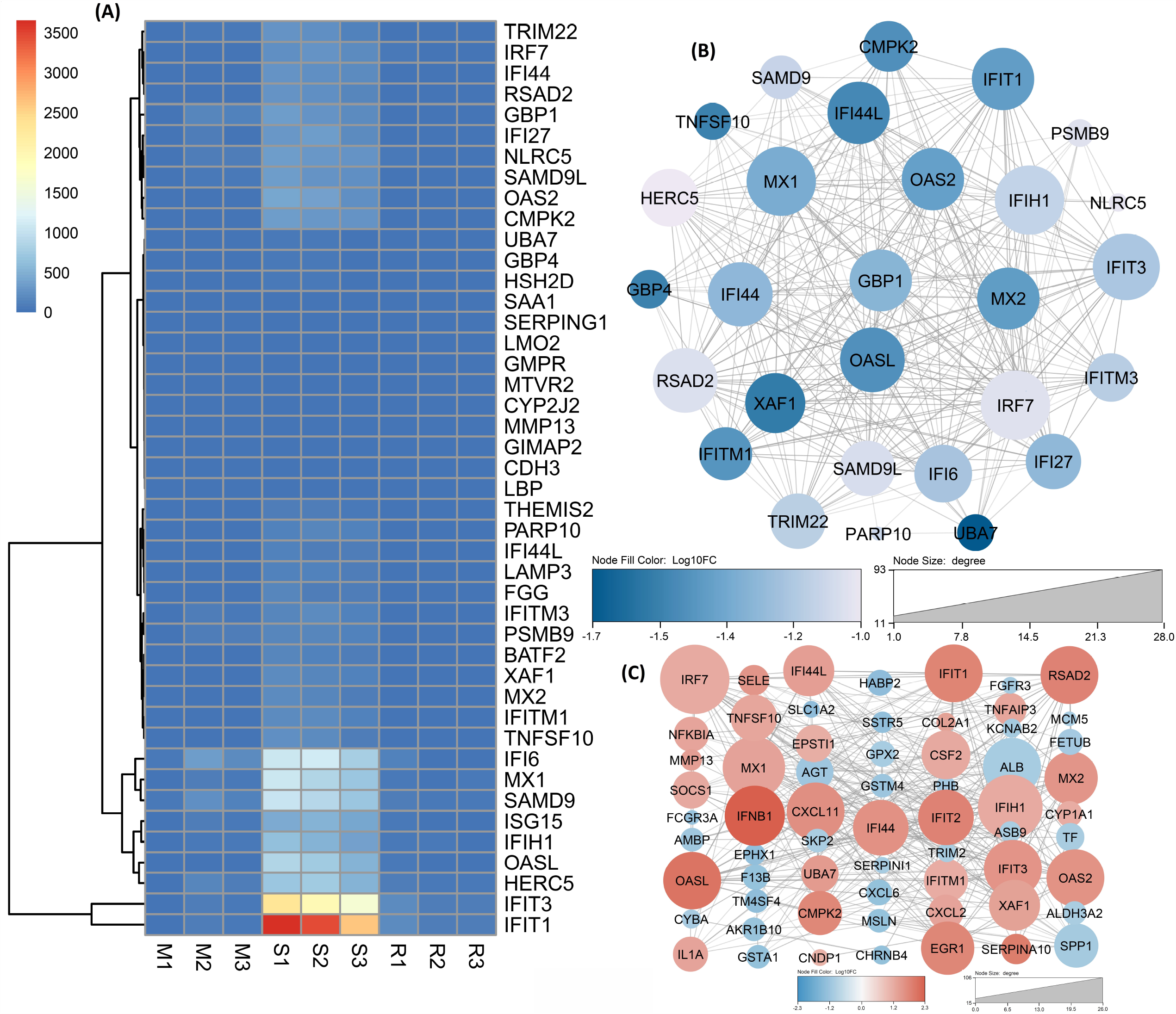
**(A)** Heatmap of normalized microarray gene expression dataset comprising triplicate samples, each of mock treated (without SARS-CoV-2 infection and ruxolitinib treatment) (M1, M2, M3), SARS-CoV-2 infected (S1, S2, S3), and SARS-CoV-2 infected treated with ruxolitinib (R1, R2, R3). The color bar denotes the microarray signal intensities, **(B)** PPI network displaying top genes with corresponding log_10_ Fold Change levels and degree connectivity values in downregulated DEGs in SARS-CoV-2 infection with and without ruxolitinib treatment, **(C)** PPI network displaying top regulatory genes with corresponding log_10_ Fold Change levels and degree connectivity values in upregulated DEGs with and without SARS-CoV-2 infection.

The Ruxolitinib treatment caused significant inhibition of reactome pathways: DDX58/IFIH1-mediated induction of interferon -I and -II signaling combined with ISG and OAS mediated antiviral response involving HERC5, UBA7, MX1, IFITM1, OAS2, IFITM3, IFIT1, GBP1, XAF1, GBP4, MX2, IFI27, OASL, TRIM22, IFIT3, RSAD2, IFI6, IRF7, NLRC5, SAA1, GBP4 and GBP1 genes (FDR ranged from 2.28E-24 to 0.0062 in reactome pathways) (Table 1). The most significant underexpressed GO process enriched in SARS-CoV-2 infected cell line with or without ruxolitinib treatment included defense response to virus (FDR 7.21E-21) among a wide array of processes (Figure 2a and Table 1). Conversely, the SARS-CoV-2 infection caused significant upregulation of similar signaling pathways and antiviral mechanism due to the action of EGR1, OASL, SOCS1, MX2, OAS2, XAF1, IFIT1, IFIT3, IFIT2, IFNB1, RSAD2, IRF7, MX1, IFITM1, IFNA5, NFKBIA, TNFAIP3 (major reactome pathways FDR ranged from 1.34E-17 to 0.0403) (Table 2). The important DEGs related to the GO process in cell line with or without SARS-CoV-2 infection (Figure 2b) were upregulated type I interferon signaling pathway (FDR 4.51E-15, log_10_FC>1) (Table 2) and downregulated homophilic cell adhesion via plasma membrane adhesion molecules (FDR 0.0299) (Table 3). The SARS-CoV-2 infection in cell line displayed significant inhibition of pathways related to cytochrome P450-mediated metabolism of drugs and xenobiotics, in addition to glutathione, ascorbate and aldarate metabolism, and chemical carcinogenesis (major KEGG pathways FDR ranged from 0.0175 to 0.0402) involving GSTA1, UGT2B15, GSTM4, UGT2B11, EPHX1, G6PD and ALDH3A2 genes (Figure 2c and Table 3).

**Table 1:**
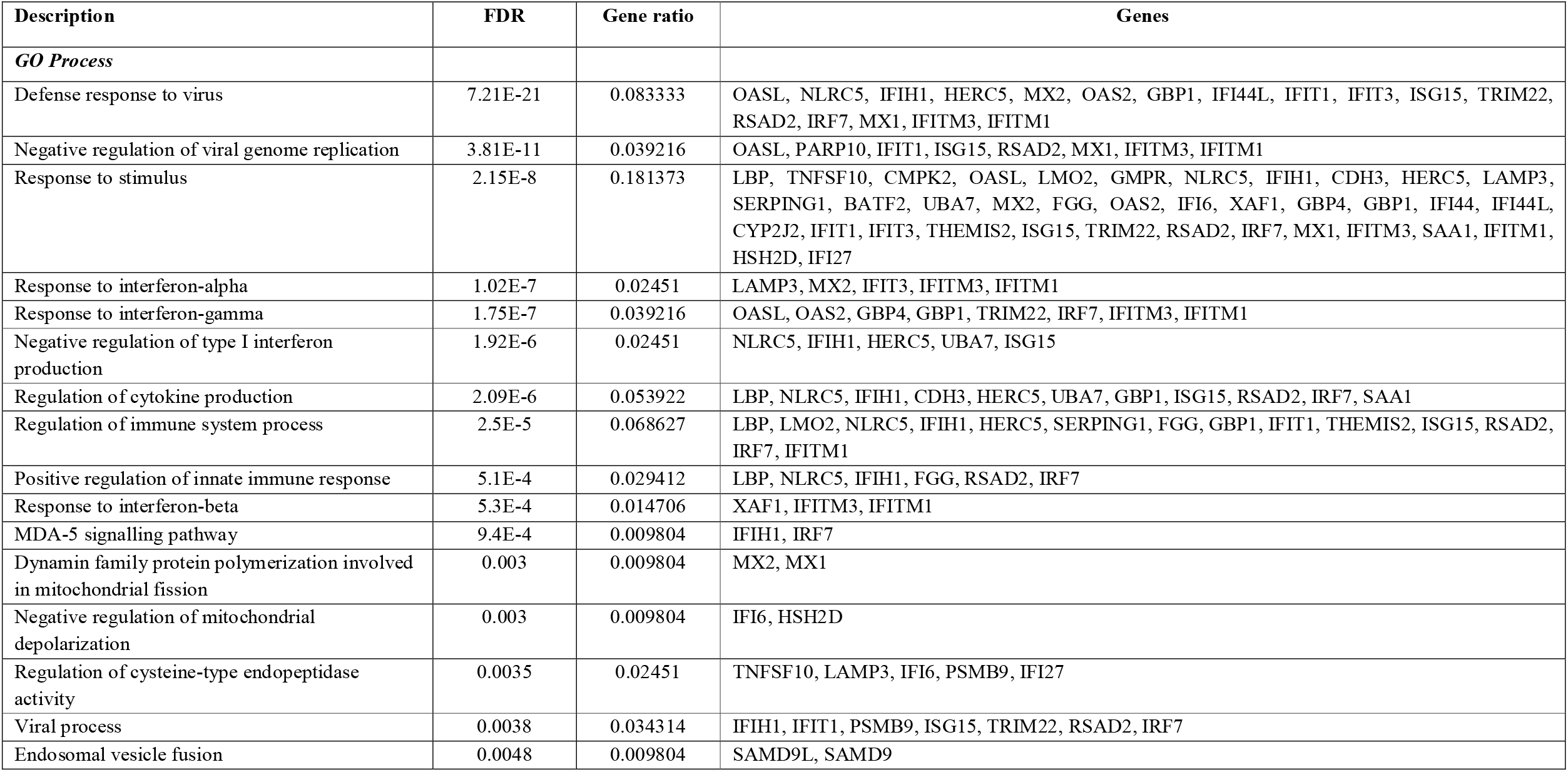
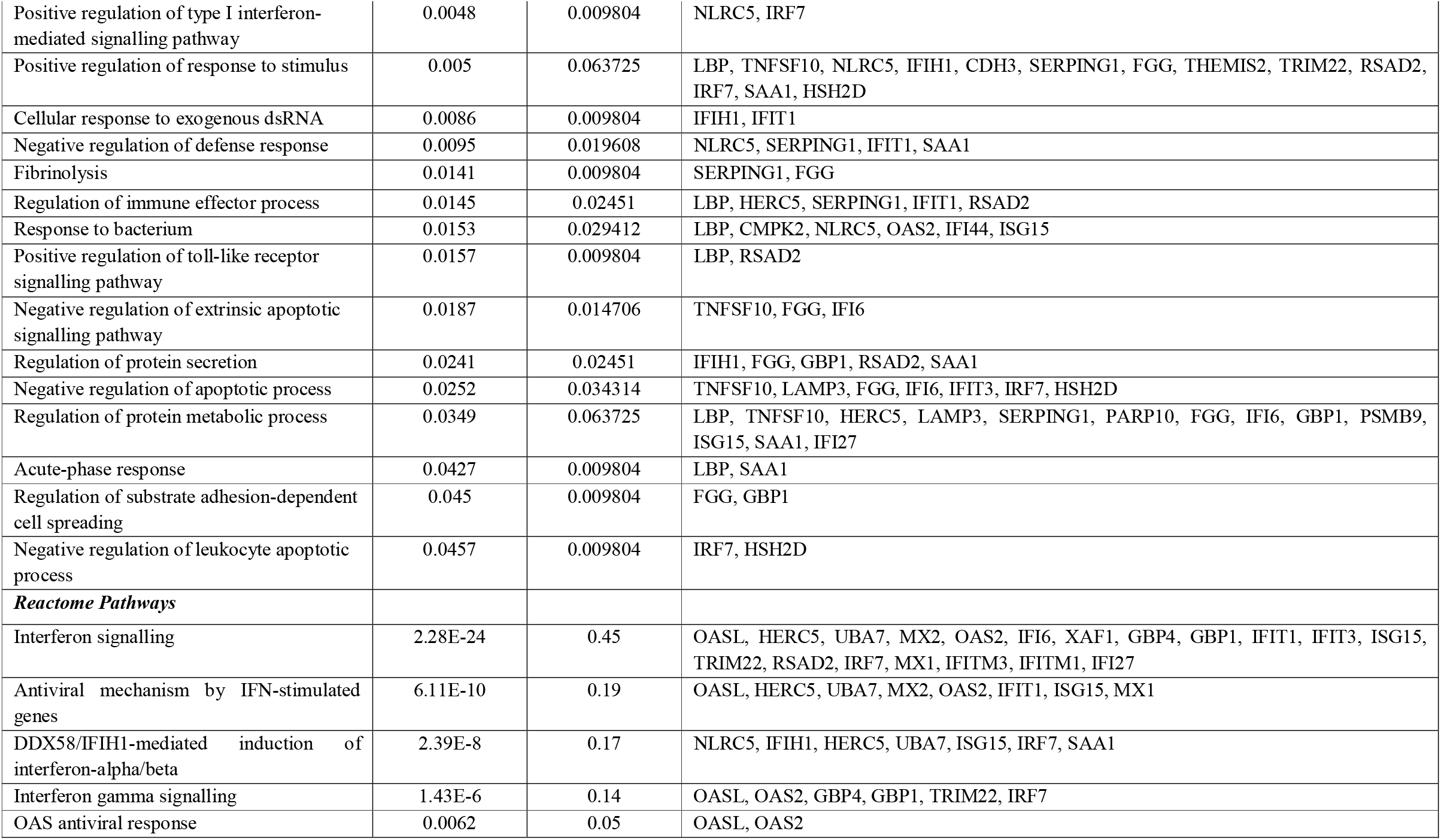
Biological process and functional enrichment of differentially expressed down regulated genes in SARS-CoV-2 infected cell line with or without ruxolitinib treatment (Protein-Protein Interaction Enrichment: 1E-16, Number of genes involved: 43, Number of Protein-Protein Interactions: 296)

**Table 2:**
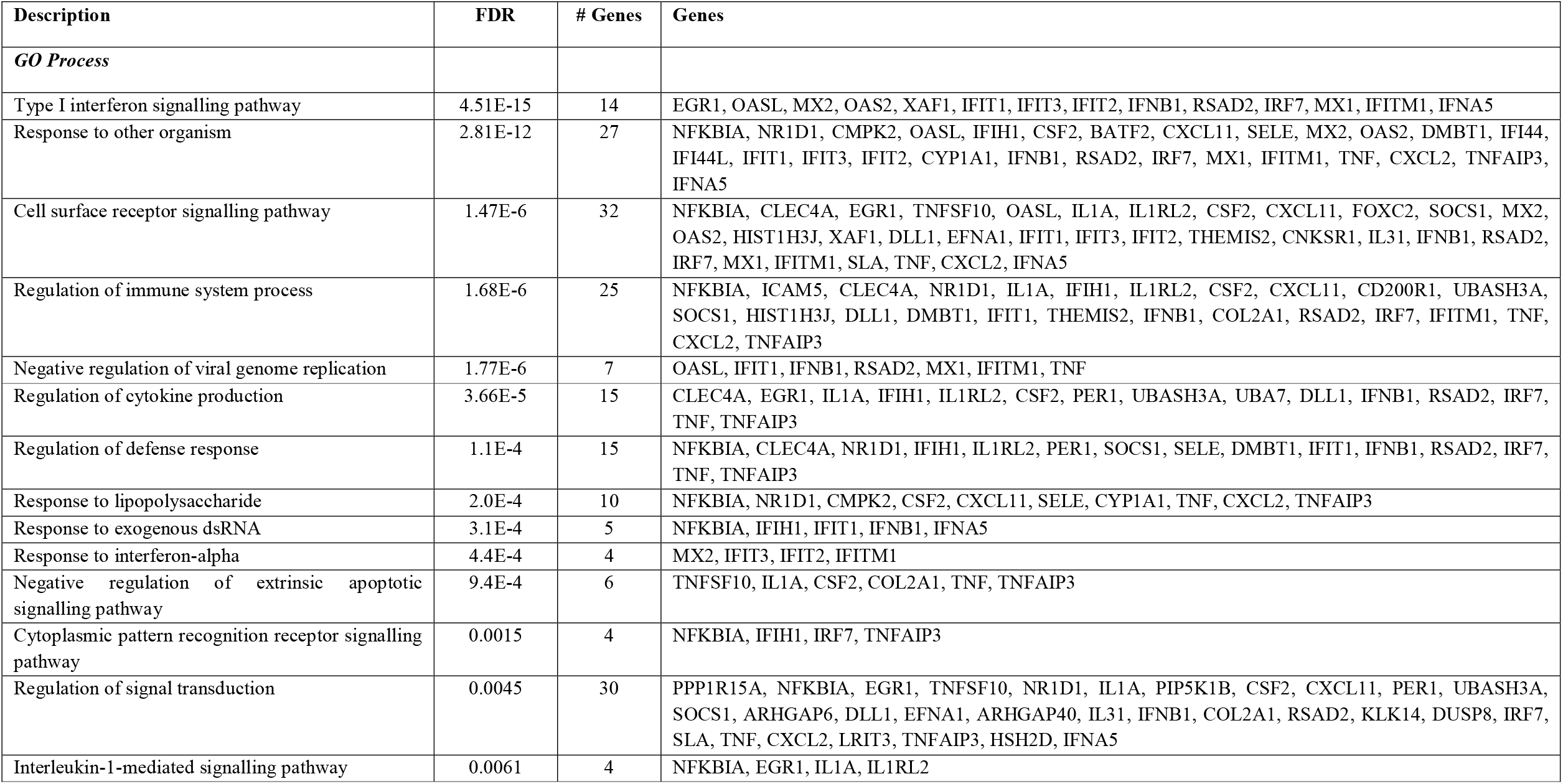
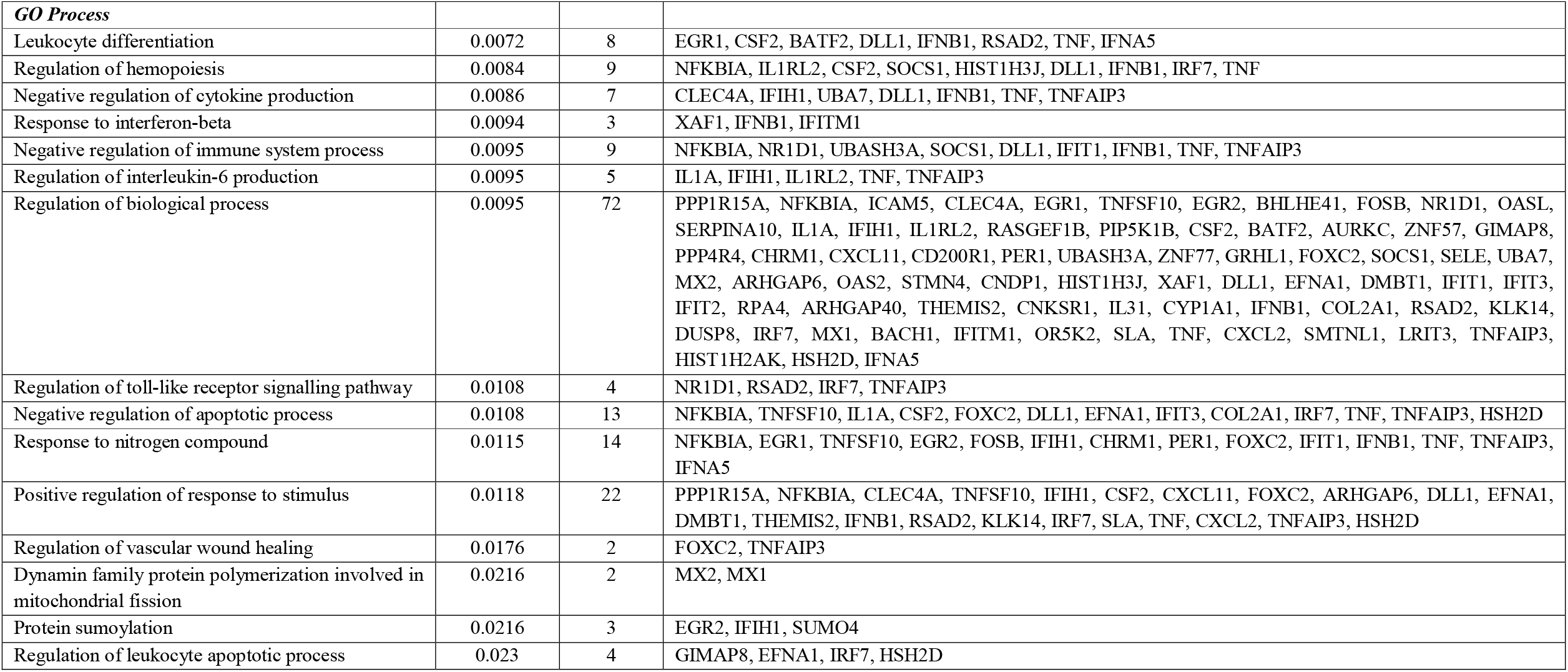
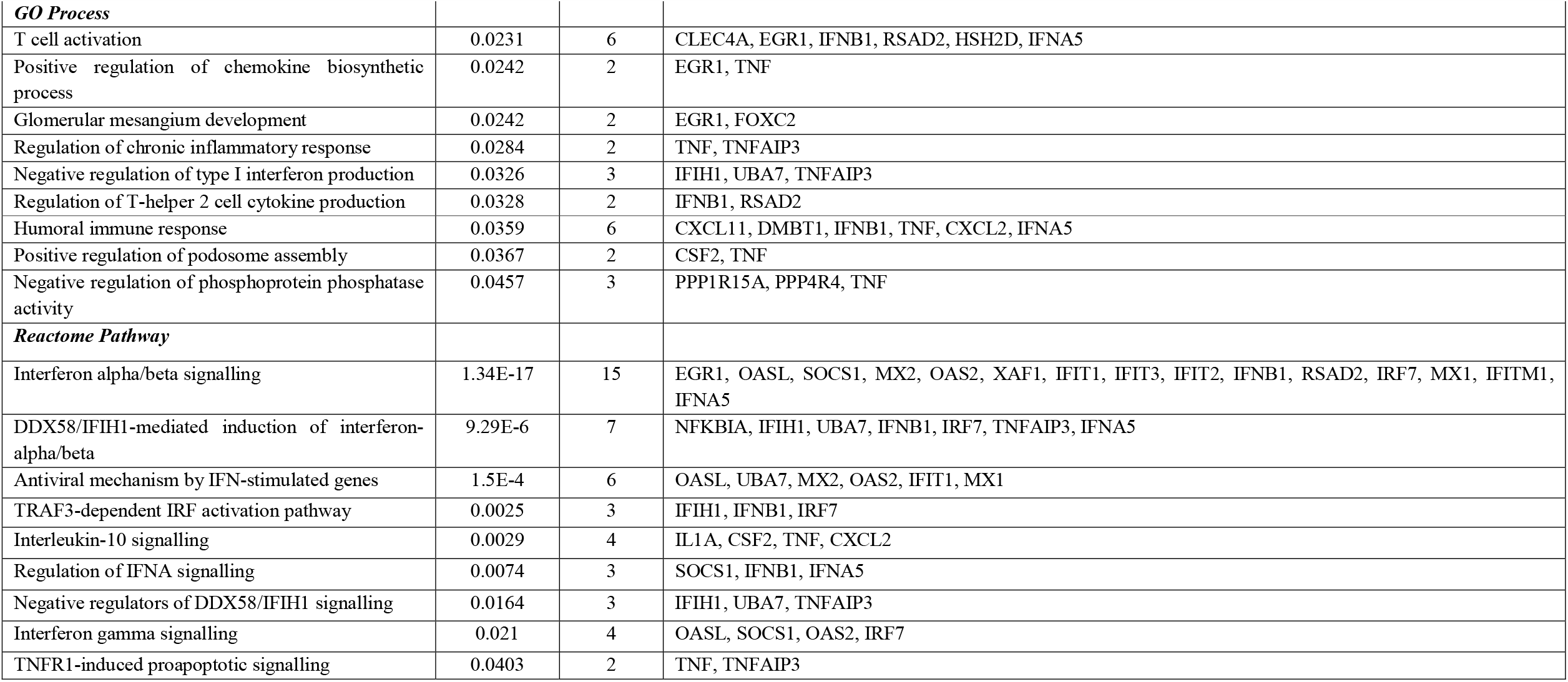
Biological process and functional enrichment of differentially expressed up regulated genes in cell line with or without SARS-CoV-2 infection (Protein-Protein Interaction Enrichment: 1E-16, Number of genes involved: 97, Number of Protein-Protein Interactions: 263)

**Table 3:**
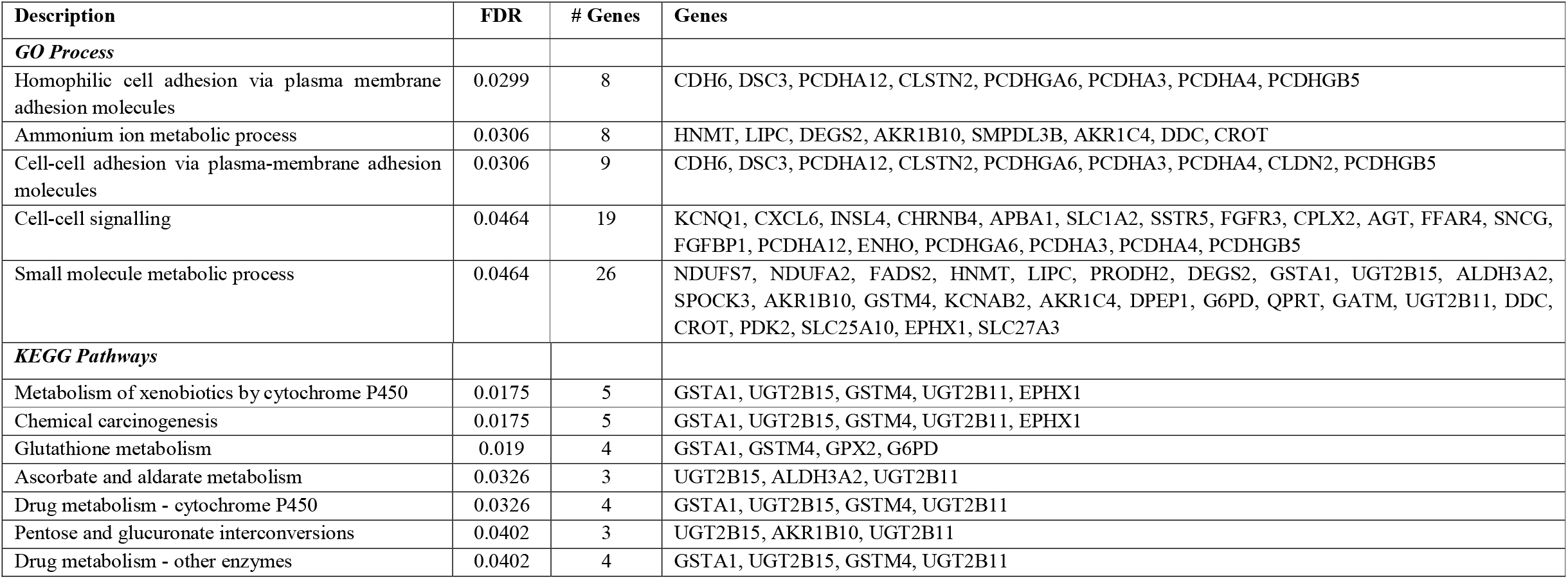
Biological process and functional enrichment of differentially expressed down regulated genes in cell line with or without SARS-CoV-2 infection (Protein-Protein Interaction Enrichment: 8.13E-9, Number of genes involved: 128, Number of Protein-Protein Interactions: 96)

**Figure 2:**
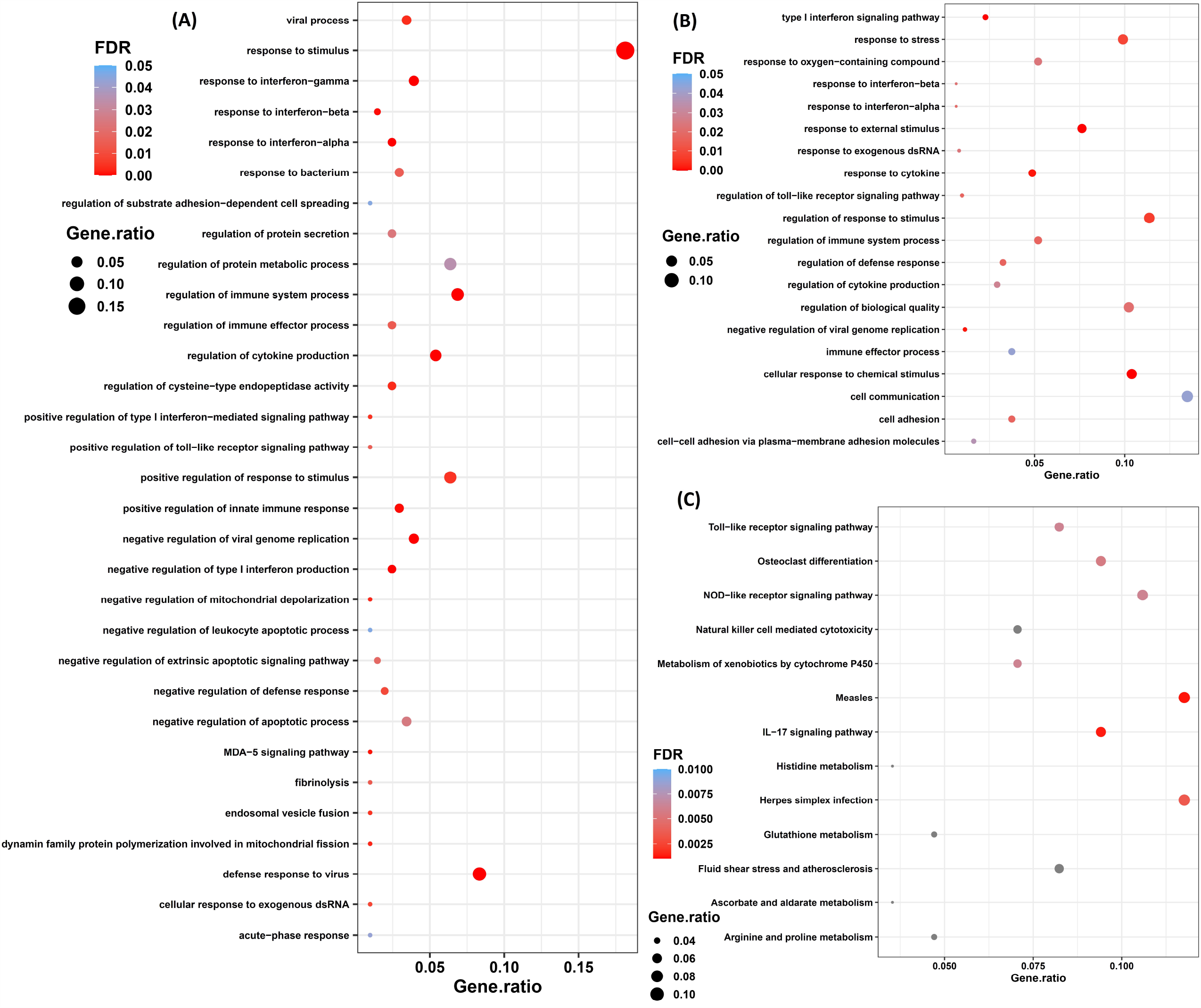
**(A)** Gene Ontology process enrichment in SARS-CoV-2 infection with and without ruxolitinib treatment, **(B)** Gene Ontology process enrichment with and without SARS-CoV-2 infection, **(C)** KEGG pathway enrichment with and without SARS-CoV-2 infection.

Host display innate immunity to SARS-CoV-2 infection primarily by IFN pathway (Table 2). The host PRRs (pattern recognition receptors) respond to the viral PAMPs (pathogen-associated molecular patterns) by eliciting RLR (RIG-I-like receptors) signaling pathway (RIG-I/DDX58, MDA5/IFIH1, NLRX1, MAVS), NOD (nucleotide-binding oligomerization domain)-like signaling pathway (NLRP3, PYCARD, CASP1), and TLR (Toll-like receptors) signaling pathway (TLR3-mediated expression of TRIF and TRAF3; TLR7-mediated expression of MyD88 and IRAK) (Figure 2c and Figure 3). Subsequent induction of downstream signaling molecules such as TBK1, IKK, NFκB and IRF result in IFN production. Activation of TYK2 and JAK1 cause STAT1/2 and IRF9 mediated stimulation of ISRE (interferon-stimulated response element)-containing promoter activating the ISGs (interferon-stimulated genes) including MxA, OAS and IFITM that cause degradation of viral RNAs and inhibit coronavirus entry (Figure 3).

**Figure 3:**
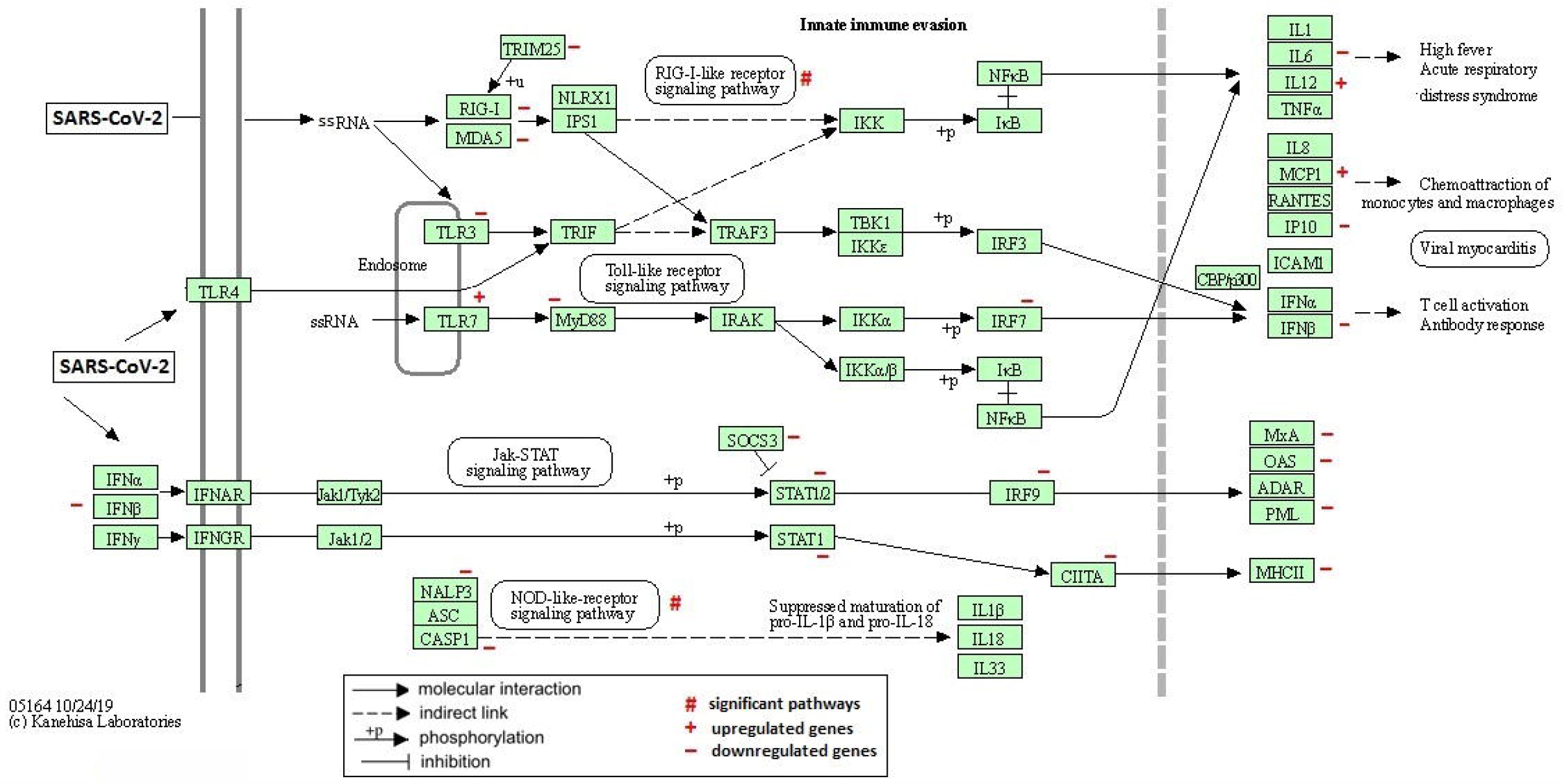
KEGG pathways of statistically enriched differentially expressed genes in innate immune evasion following SARS-CoV-2 infection and ensuing events.

Overall, together the Type-I/III interferon production and the resultant signaling are vital in regulating early SARS-CoV-2 infection. SARS-CoV-2 evade host innate immune response by expressing several IFN antagonists (Yuen et al. 2020). According to the current investigation with GSE147507 dataset, ruxolitinib treatment exhibited beneficiary function by dysregulating IFN antagonists achieving the normal cellular function and suppression of viral pathogenesis. Inhibition of IFN antagonists nsp13 (upregulation of DDX5, down regulation of RNGTT), nsp14 (decrease of RNMT), nsp15 (downregulation), nsp3 (inhibition of UBE2N mediated DUB activity) were detected. In addition, other IFN antagonists include orf3b, orf6, M, N, nsp1, nsp7 and nsp16 causing suppression of interferon production and signaling (Yuen et al. 2020). In context to the safety concerns, ruxolitinib was mostly well tolerated with some reports of toxicity including myelosuppression resulting in dose-limiting thrombocytopenia and anemia, and viral reactivations (Ajayi et al. 2018). Further, the CYP3A4-mediated ruxolitinib metabolism is required to be monitored mainly when co-administration with strong CYP3A4 inhibitors (Ajayi et al. 2018). However, no direct correlation between viral load and ruxolitinib treatment was found considering that the interval between the appearance of COVID-19 symptoms and the commencement of ruxolitinib treatment was a median of 9 days possibly when viral load subsided with beginning of hyperinflammation damage (Pan et al. 2020).

The expression of genes related to severe COVID-19 features including lymphopenia, higher concentrations of inflammatory cytokines, pyroptosis and ARDS, were reversed with ruxolitinib treatment, as found in the present study (Table 4). The aberrant immune activation as an antiviral response leading to overproduction of cytokines were substantially reduced as evident from the suppression of genes related to interferon I signalling as well as anti-viral and proinflammatory ISGs. Although reversed, there was not enough evidence of significant gene expression achieving the normal circulating levels of monocytes, macrophages, and eosinophils suppressed due to SARS-CoV-2 infection (Table 4).

**Table 4:**
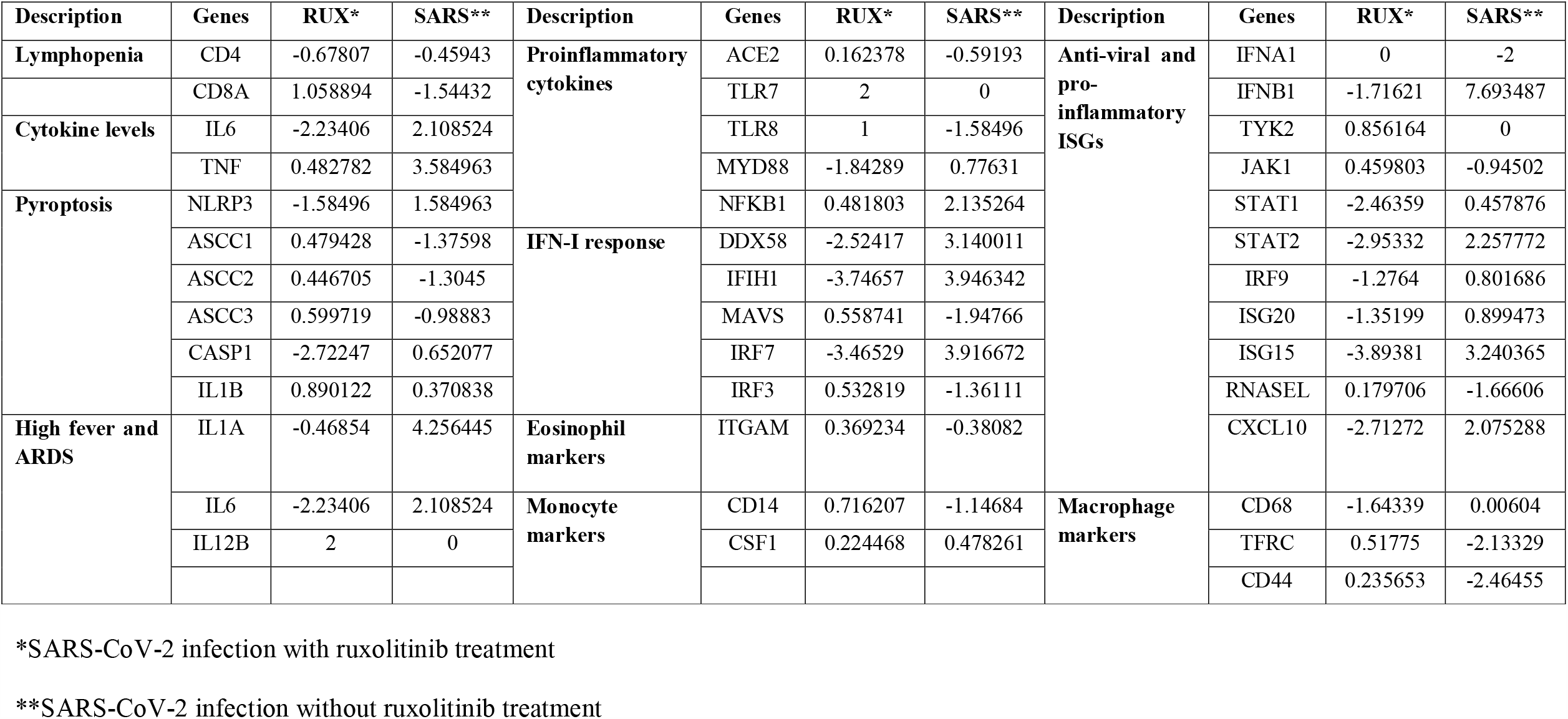
Log_10_FoldChange gene expression of COVID-19 features in SARS-CoV-2 infected cell line with or without ruxolitinib treatment

### Conclusion

The present study indicated that ruxolitinib treatment elicited similar response equivalent to that of SARS-CoV-2 uninfected situation by causing stimulation of defense response in host against virus infection by RLR and NOD like receptor signaling pathways. Further, the effect of ruxolitinib in SARS-CoV-2 infection was mainly caused by significant suppression of IFIH1, IRF7 and MX1 genes as well as inhibition of DDX58/IFIH1-mediated induction of interferon-I and -II signaling combined with ISG and OAS mediated antiviral response. It is expected that an exploration into the mechanism of agents possessing any possibility of reducing COVID-19 related cytokine storm will contribute to the scientific understanding of this complex disease, and might serve as a treatment option in life threatening conditions like respiratory insufficiency caused by COVID-19 associated ARDS, or to find avenues for the improvement of patient outcome in COVID-19 in whom respiratory symptoms are deteriorating nonetheless with still reversible lung impairment.

## Data Availability

Source of the data analyzed are mentioned in the article, and the data derived in the current study are with the authors of this article.

## Author’s contribution

Both MM and SM designed the study, analysed and interpreted data, discussed and wrote the manuscript.

## Funding Source

Nil

## Declaration of competing interests

There is no conflict of interest by the authors.

## Notes

### Competing Interest Statement

The authors have declared no competing interest.

### Clinical Trial

nil

### Funding Statement

No funding source is relevant to this study.

### Author Declarations

Details are given in the manuscript related to analysis of publicly available dataset.

